# COVID-19 Disease Severity and Associated Factors among Ethiopian Patients: A study of the Millennium COVID-19 Care Center

**DOI:** 10.1101/2020.10.09.20209999

**Authors:** Tigist W. Leulseged, Kindalem G. Abebe, Ishmael S. Hassen, Endalkachew H. Maru, Wuletaw C. Zewde, Negat W. Chamiso, Kalkidan T. Yegele, Abdi B. Bayisa, Dagne F. siyoum, Mesay G. Edo, Edmialem G. Mesfin, Meskerem N. Derejie, Helina K. Shiferaw

## Abstract

**Background:** The COVID-19 pandemic started a little later in Ethiopia than Europe and most of the initial cases were reported to have a milder disease course and a favorable outcome. This changed as the disease spread into the population and the more vulnerable began to develop severe disease. Understanding the risk factors for severe disease in Ethiopia was needed to provide optimal health care services in a resource limited setting.

**Objective:** The study assessed COVID-19 patients admitted to Millennium COVID-19 Care Center in Ethiopia for characteristics associated with COVID-19 disease severity.

**Methods:** A cross-sectional study was conducted from June to August 2020 among 686 randomly selected 686 patients. Chi-square test was used to detect the presence of a statistically significant difference in the characteristics of the patients based on disease severity (Mild vs Moderate vs Severe). A multinomial logistic regression model was used to identify risk factors of COVID-19 disease severity where Adjusted Odds ratio (AOR), 95% CIs for AOR and P-values were used for significance testing.

**Results:** Having moderate as compared with mild disease was significantly associated with having hypertension *(AOR=2.30, 95%CI=1.27,4.18, p-value=0.006), diabetes mellitus (AOR=2.61, 95%CI=1.31,5.19, p-value=0.007 for diabetes mellitus), fever (AOR=6.12, 95%CI=2.94,12.72, p-value=0.0001)* and headache *(AOR=2.69, 95%CI=1.39,5.22, p-value=0.003)*. Similarly, having severe disease as compared with mild disease was associated with age group *(AOR=4.43, 95%CI=2.49,7.85, p-value=0.0001 for 40-59 years and AOR=18.07, 95%CI=9.29,35.14, p-value=0.0001 for ≥ 60 years)*, sex (*AOR=1.84, 95%CI=1.12,3.03, p-value=0.016)*, hypertension *(AOR=1.97, 95%CI=1.08,3.59, p-value=0.028)*, diabetes mellitus *(AOR=3.93, 95%CI=1.96,7.85, p-value=0.0001)*, fever *(AOR=13.22, 95%CI=6.11, 28.60, p-value=0.0001)* and headache *(AOR=4.82, 95%CI=2.32, 9.98, p-value=0.0001)*. In addition, risk factors of severe disease as compared with moderate disease were found to be significantly associated with age group *(AOR=4.87, 95%CI=2.85, 8.32, p-value=0.0001 for 40-59 years and AOR=18.91, 95%CI=9.84,36.33, p-value=0.0001 for* ≥ *60 years)*, fever *(AOR=2.16, 95%CI=1.29,3.63, p-value=0.004)* and headache *(AOR=1.79, 95%CI=1.03, 3.11, p-value=0.039)*.

**Conclusions:** Risk factors associated with severe COVID-19 in Ethiopia are being greater than 60 years old, male, a diagnosis of hypertension, and diabetes mellitus, and the presence of fever and headache. This is consistent with severity indicators identified by WHO and suggests the initial finding of milder disease in Ethiopia may have been because the first people to get COVID-19 in Ethiopia were less than 60 years of age with fewer health problems.

## Introduction

The Coronavirus Infectious Disease 2019 (COVID-19) caused by the new coronavirus Severe Acute Respiratory Syndrome Coronavirus Type 2 (SARS-CoV-2) has affected the entire world resulting in loss of millions of lives and causing a significant burden on the health care system and the economy in general [1]. According to the World Health Organization (WHO) daily situation report, the first infected case was identified in Africa a little later than the rest of the world after it has already caused a significant morbidity and mortality in others.

In Ethiopia, the first case of COVID-19 was diagnosed two days after the WHO has declared the disease to be a pandemic on March 11, 2020. According to the Ethiopian Federal Ministry of Health Daily COVID-19 report, As of October 2, 2020, there were a total of 44, 101 active cases with 296 severe patients [2]. As can be seen, majority of the reported cases have milder disease presentation with favorable outcome.

Studies show that severe disease seems to be determined by socio-demographic characteristics including male sex and older age. Having a history of pre-existing co-morbid illness particularly hypertension, diabetes, severe asthma, cancer, renal disease, cardiovascular, cerebrovascular diseases, and other co-morbidities were also found to be predictors of severe disease [3-11].

Disease severity is also reported to be associated with lower oxygen saturation and abnormal laboratory markers including higher levels of leukocyte count, neutrophil count, high sensitivity C reactive protein, procalcitonin, ferritin, interleukin 2,6 and 8 receptors, tumor necrosis factor α, D-dimer, fibrinogen, lactate dehydrogenase, N-terminal pro-brain natriuretic peptide, cytokine, LDH and lower levels of CD4 count and deranged lymphocyte count [9, 12-14].

With the differing pattern in disease presentation, progression and outcome in Africa in general as compared with the rest of the world as observed so far and also considering the existing endemic disease pattern, conducting a study in Africa is crucial to understand the disease pattern in the local context. Because knowing risk factors of developing severe disease is important as severe disease is associated with worse outcome so that stratified and focused patient management and preventive practices can be provided which is a practical solution in a resource limited country like Ethiopia where the health care infrastructure cannot meet the potentially growing demand of intensive care unit admission

Therefore, the objective of this study was to identify the risk factors of disease severity among COVID-19 patients admitted to Millennium COVID-19 Care Center in Addis Ababa, Ethiopia.

## Materials and Methods

### Study Design, Setting and Population

An institution-based cross-sectional study was conducted at Millennium COVID-19 Care Center (MCCC), a 1000 bed makeshift hospital in Addis Ababa, Ethiopia dedicated for isolating and treating COVID-19 cases. The center is remodeled from the previous Millennium hall/ Addis park which was a multipurpose recreational, meeting and exhibition center. It is the first and also the largest center in the country with majority of the cases in the country admitted to the Center especially during the first few months of the pandemic in the country before the other COVID-19 wings in permanent hospitals and regional Centers started function. Even after that, it remained to be a center with the largest admission. At the beginning, since there were few cases, the MCCC and also other Centers in the country were used as both a quarantine and treatment center in order to halt the transmission of the disease. Therefore, anyone who tested positive for SARS-Cov-2 gets admitted to COVID-19 Centers despite the age, disease severity, presence of symptoms and co-morbidity status.

The source population was all patients admitted to MCCC with a confirmed diagnosis of COVID-19 using RT-PCR from June to August 2020.

The study population was all selected COVID-19 patients who were on treatment and follow up at MCCC during the three month period and who fulfill the inclusion criteria.

### Sample Size Determination and Sampling Technique

The sample size to identify risk factors of disease severity was determined using a double population proportion formula with the assumptions of; 95% confidence interval, power of 80%, the proportion of males who had severe disease as 0.80, proportion of females who had non-severe disease as 0.75 and considering a non-response rate of 10%. Therefore, the total sample size calculated becomes 689 COVID-19 patients. A simple random sampling method using table of random numbers was used to select the study participants.

### Eligibility criteria

All selected COVID-19 patients who were admitted to MCCC during the three months follow-up period and who consented to participate were included in the study.

### Operational Definitions

**COVID-19 disease** [15]:

– **Mild Disease:** characterized by fever, malaise, cough, upper respiratory symptoms, and/or less common features of COVID-19 (headache, loss of taste or smell etc…)
– **Moderate Disease:** Patients with lower respiratory symptom/s. They may have infiltrates on chest X-ray. These patients are able to maintain oxygenation on room air.
– **Severe COVID-19 disease:** Includes patients who have developed complications. The following features can define severe illness.
  - Hypoxia: SPO2 ≤ 93% on atmospheric air or PaO2:FiO2 < 300mmHg (SF ratio < 315)
  - Tachypnea: in respiratory distress or RR>30 breaths/minutes
  - More than 50% involvement seen on chest imaging

### Data Collection Procedures and Quality Assurance

An interviewer-administered questionnaire that consists of the variables of interest was developed from the patient registration and follow up form which was adopted from the WHO patient admission and management guideline and used at the center for patient follow up.

The data collection tool was pretested on 35 randomly selected patients and their medical charts which were not included in the final data collection and necessary amendment on the data collection tool was made.

Training on the basics of the questionnaire and data collection technique was given for ten data collectors (BSc nurses and General practitioners) and two supervisors (General practitioner and public health specialist).

Data consistency and completeness was checked before an attempt was made to enter the code and analyze the data.

### Statistical Analysis

The extracted data were coded, entered into Epi-Info version 7.2.1.0, cleaned, stored, and exported to SPSS version 23.0 software for analysis. Categorical covariates were summarized using frequencies and percentages and numerical variables were summarized with a mean value. A Chi-square test/ Fischer’s exact test was run to compare the underlying characteristics of the patients based on disease severity. A statistically significant difference was detected for variables with a P-value of ≤ 0.05. The presence of multi-collinearity was checked for the independent variables fit on the final model and the VIF result ranges from 1.081 to 1.314 showing that there is no multi-collinearity issue in the final mode.

The association between disease severity and determinant variables were analyzed using Multinomial Logistic Regression. Univariate analysis was done at 25% level of significance to screen out independent variables used in the multivariable multinomial Logistic regression model. The adequacy of the final model was assessed using goodness of fit test and the final model fitted the data well (Pearson x ^2^ _(118)_ =141.005, p-value = 0.073 and Deviance x^2^_(118)_ =134.542, p-value = 0.142). For the multinomial Logistic regression, 95% confidence interval for AOR was calculated and variables with p-value ≤ 0.05 were considered as statistically associated with COVID-19 disease severity at admission.

### Ethics approval

The study was conducted after obtaining ethical clearance from St. Paul’s Hospital Millennium Medical College Institutional Review Board. After assessing their level of consciousness, written informed consent was obtained from the participants / parents or legal guardians for minors. The study had no risk/negative consequence on those who participated in the study. Medical record numbers were used for data collection and personal identifiers were not used in the research report. Access to the collected information was limited to the principal investigator and confidentiality was maintained throughout the project.

## Result

### Socio-demographic, co-morbid illness and disease related characteristics and comparison based on disease severity

From the 689 samples selected, information was collected from 686 patients making the response rate 99.5%. The median age of the participants was 40.0 (IQR, 30.0-57.3) years. The majority (63.1%) were males. Two hundred sixty-seven (38.9%) of the patients had a history of one or more co-morbid illnesses. The most common co-morbid illness in the study population was hypertension (21.1%), followed by diabetes mellitus (16.6%), cardiac disease (5.2%) and Asthma (4.7%), tuberculosis (1.7%). Other co-morbid illnesses like chronic kidney disease and neurologic disorder constituted less than 2% of the co-morbid illnesses reported. Twenty-five (3.6%) of the patients were Khat chewers and nine (1.3%) were smokers.

A statistically significant difference in disease severity was found among the different groups of patients by age group, sex, co-morbid illness, hypertension, and diabetes mellitus. A significant proportion of patients 40 to 59 years (29.4% Vs 29.9% Vs 40.6%, p-value=0.0001) and ≥ 60 years (16.6% Vs 16.6% Vs 66.9%, p-value=0.0001) had severe disease and the younger age group of < 40 years had mild disease (46.2% Vs 41.1% Vs 12.7%, p-value=0.0001). Based on sex, a significantly higher proportion of female patients had mild disease (41.1% Vs 31.2% Vs 27.7%, p-value=0.021). On the contrary, males had severe disease (31.2% Vs 33.3% Vs 35.6%, p-value=0.021). A significantly higher proportion of patients with one or more co-morbid illnesses had severe disease followed by moderate and then mild disease (20.6% Vs 30.3% Vs 49.1%, p-value=0.0001). Similarly, patients with hypertension (17.2% Vs 29.7% Vs 53.1%, p-value=0.0001) and diabetes (12.3% Vs 28.9% Vs 58.8%, p-value=0.0001) had severe disease followed by moderate and then mild disease. (**Table 1**)

**Table 1:** Socio-demographic, co-morbid illness, disease related characteristics and comparison based on disease severity among patients (n=686)

| **Variable** | **Mild**<br>**(n=239)**<br>**(%)** | **Moderate**<br>**(n=223) (%)** | **Severe**<br>**(n=224) (%)** | **Total (%)** | **P-value** |
| --- | --- | --- | --- | --- | --- |
| **Age** |  |  |  |  |  |
| < 40 | 156 (65.3) | 139 (62.3) | 43 (19.2) | 338 (49.3) | **0.0001\*** |
| 40-59 | 58 (24.3) | 59 (26.5) | 80 (35.7) | 197 (28.7) |  |
| ≥ 60 | 25 (10.4) | 25 (11.2) | 101 (45.1) | 151 (22.0) |  |
| **Sex** |  |  |  |  |  |
| Female | 104 (43.5) | 79 (35.4) | 70 (31.2) | 253 (36.9) | **0.021\*** |
| Male | 135 (56.5) | 144 (64.6) | 154 (68.8) | 433 (63.1) |  |
| **Co-morbid illness** |  |  |  |  |  |
| Yes | 55 (23.0) | 81 (36.3) | 131 (58.5) | 267 (38.9) | **0.0001\*** |
| No | 184 (76.9) | 142 (63.7) | 93 (41.5) | 419 (61.1) |  |
| **Hypertension** |  |  |  |  |  |
| Yes | 25 (10.5) | 43 (19.2) | 77 (34.4) | 145 (21.1) | **0.0001\*** |
| No | 214 (89.5) | 180 (80.7) | 147 (65.6) | 541 (78.8) |  |
| **Diabetes Mellitus** |  |  |  |  |  |
| Yes | 14 (5.9) | 33 (14.8) | 67 (29.9) | 114 (16.6) | **0.0001\*** |
| No | 225 (94.1) | 190 (85.2) | 157 (70.1) | 572 (83.3) |  |
| **Asthma** |  |  |  |  |  |
| Yes | 9 (3.8) | 9 (4.0) | 14 (6.3) | 32 (4.7) | 0.387 |
| No | 230 (96.2) | 214 (95.9) | 210 (93.8) | 654 (95.3) |  |
| **Khat chewing** |  |  |  |  |  |
| Yes | 5 (2.1) | 8 (3.6) | 12 (5.4) | 25 (3.6) | 0.173 |
| No | 234 (97.9) | 215 (96.4) | 212 (94.6) | 661 (96.4) |  |

### Presenting symptom related characteristics and comparison based on disease severity

The most common presenting symptom was cough (56.4%) followed by shortness of breath (26.9%), fatigue (23.2%), fever (20.9%), headache (16.5%), chest pain (16.1%), sore throat (13.7%), arthralgia (11.2%), myalgia (10.1%), and runny nose (5.1%).

As shown in Table 2, the chi-square test result shows that a significantly higher proportion of patients with any of the above presenting symptoms had sever disease, followed by moderate and then mild disease. (**Table 2**)

**Table 2:** Presenting symptom related characteristics and comparison based on disease severity among COVID-19 patients (n=686)

| **Variables** | **Mild**<br>**(n=239)**<br>**(%)** | **Moderate**<br>**(n=223) (%)** | **Severe**<br>**(n=224)**<br>**(%)** | **Total (%)** | **P-value** |
| --- | --- | --- | --- | --- | --- |
| **Fever** |  |  |  |  |  |
| Yes | 10 (4.2) | 53 (23.8) | 81 (36.2) | 144 (20.9) | **0.0001\*** |
| No | 229 (95.8) | 170 (76.2) | 143 (63.8) | 542 (79.1) |  |
| **Sore throat** |  |  |  |  |  |
| Yes | 14 (5.9) | 40 (17.9) | 40 (17.9) | 94 (13.7) | **0.0001\*** |
| No | 225 (94.1) | 183 (82.1) | 184 (82.1) | 592 (86.3) |  |
| **Runny nose** |  |  |  |  |  |
| Yes | 3 (1.3) | 16 (7.2) | 16 (7.1) | 35 (5.1) | **0.004\*** |
| No | 236 (98.7) | 207 (92.8) | 208 (92.9) | 651 (94.9) |  |
| **Myalgia** |  |  |  |  |  |
| Yes | 7 (2.9) | 25 (11.2) | 37 (16.5) | 69 (10.1) | **0.0001\*** |
| No | 232 (97.1) | 198 (88.8) | 187 (83.4) | 617 (89.9) |  |
| **Arthralgia** |  |  |  |  |  |
| Yes | 9 (3.8) | 23 (10.3) | 45 (20.1) | 77 (11.2) | **0.0001\*** |
| No | 230 (96.2) | 200 (89.7) | 179 (79.9) | 609 (88.8) |  |
| **Fatigue** |  |  |  |  |  |
| Yes | 11 (4.6) | 42 (18.8) | 106 (47.3) | 159 (23.2) | **0.0001\*** |
| No | 228 (95.4) | 181 (81.2) | 118 (52.7) | 527 (76.8) |  |
| **Headache** |  |  |  |  |  |
| Yes | 15 (6.3) | 42 (18.8) | 56 (25.0) | 113 (16.5) | **0.0001\*** |
| No | 224 (93.7) | 181 (81.2) | 168 (75.0) | 573 (83.5) |  |

### Results of risk factors of COVID-19 disease severity (Mild Vs Moderate Vs Severe)

Based on the result of the Univariate analysis at 25% level of significance; Age group, sex, hypertension, diabetes mellitus, fever, and headache were found to be significantly associated with COVID-19 disease severity (Mild Vs Moderate Vs Severe).

On the multivariable multinomial logistic regression at 5% level of significance; age group, hypertension, diabetes mellitus, fever, and headache were found to be significantly associated with COVID-19 disease severity. Accordingly, after adjusting for other covariates, for patients in the age range of 40 to 59 years and ≥ 60 years, the odds of having severe disease as compared with mild disease were 4.428 and 18.070 times than patients < 40 years, respectively (AOR= 4.43, 95% CI= 2.49, 7.85, p-value=0.0001 for 40-59 years and AOR=18.07, 95% CI= 9.29, 35.14, p-value=0.0001 for ≥ 60 years). Similarly, the odds of having severe disease as compared with moderate disease among patients 40 to 59 years and ≥ 60 years were 4.87 and 18.906 times than patients < 40 years, respectively (AOR= 4.87, 95% CI= 2.85, 8.32, p-value=0.0001 for 40-59 years and AOR= 18.91, 95% CI= 9.84, 36.33, p-value=0.0001 for ≥ 60 years). But age group didn’t show a significant association with disease severity between moderate and mild cases.

A significant association of sex with disease severity was found only between severe cases as compared with mild. Being male was associated with a 1.84 odds of having severe disease as compared with mild disease than females (AOR=1.84, 95% CI=1.12, 3.03, p-value=0.016).

Regarding co-morbid illness, having hypertension and diabetes was significantly associated with disease severity between moderate Vs mild and moderate Vs severe disease, but not between severe Vs moderate disease. For patients with hypertension and diabetes, the odds of having moderate disease as compared with mild disease were 2.30 and 2.61 times compared to patients with no such illnesses, respectively (AOR= 2.30, 95% CI= 1.27, 4.18, p-value=0.006 for hypertension and AOR= 2.61, 95% CI= 1.31, 5.19, p-value=0.007 for diabetes mellitus). Similarly, the odds of having severe disease as compared with mild disease for hypertensive and diabetic patients were 1.97 and 3.93 times patients with no such illnesses, respectively (AOR= 1.97, 95% CI= 1.08, 3.59, p-value=0.028 for hypertension and AOR= 3.93, 95% CI= 1.96, 7.85, p-value=0.0001 for diabetes mellitus).

Concerning presenting symptom, having fever and headache was significantly associated with disease severity; moderate Vs mild, severe Vs mild, and severe Vs moderate. The odds of having moderate disease as compared with mild disease for patients with fever and headache were 6.12 and 2.69 times than patients with no such symptoms, respectively (AOR= 6.12, 95% CI= 2.94, 12.72, p-value=0.0001 for fever; AOR= 2.69, 95% CI= 1.39, 5.22, p-value=0.003 for headache). The odds of having severe disease as compared with mild disease for patients with fever and headache were 13.22 and 4.82 times than patients with no such symptoms, respectively (AOR= 13.22, 95% CI= 6.11, 28.60, p-value=0.0001 for fever; AOR= 4.82, 95% CI= 2.32, 9.97, p-value=0.0001 for headache). Similarly, the odds of having severe disease as compared with moderate disease for patients with fever and headache were 2.16 and 1.79 times compared to patients with no such symptoms, respectively (AOR= 2.16, 95% CI= 1.29, 3.63, p-value=0.004 for fever; AOR= 1.79, 95% CI= 1.03 3.11, p-value=0.039 for headache). (**Table 3**)

**Table 3:**
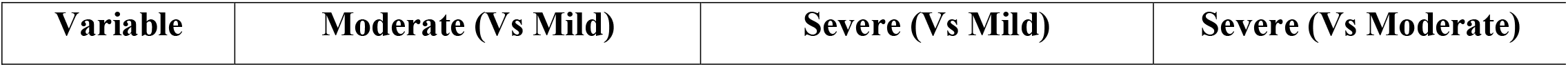
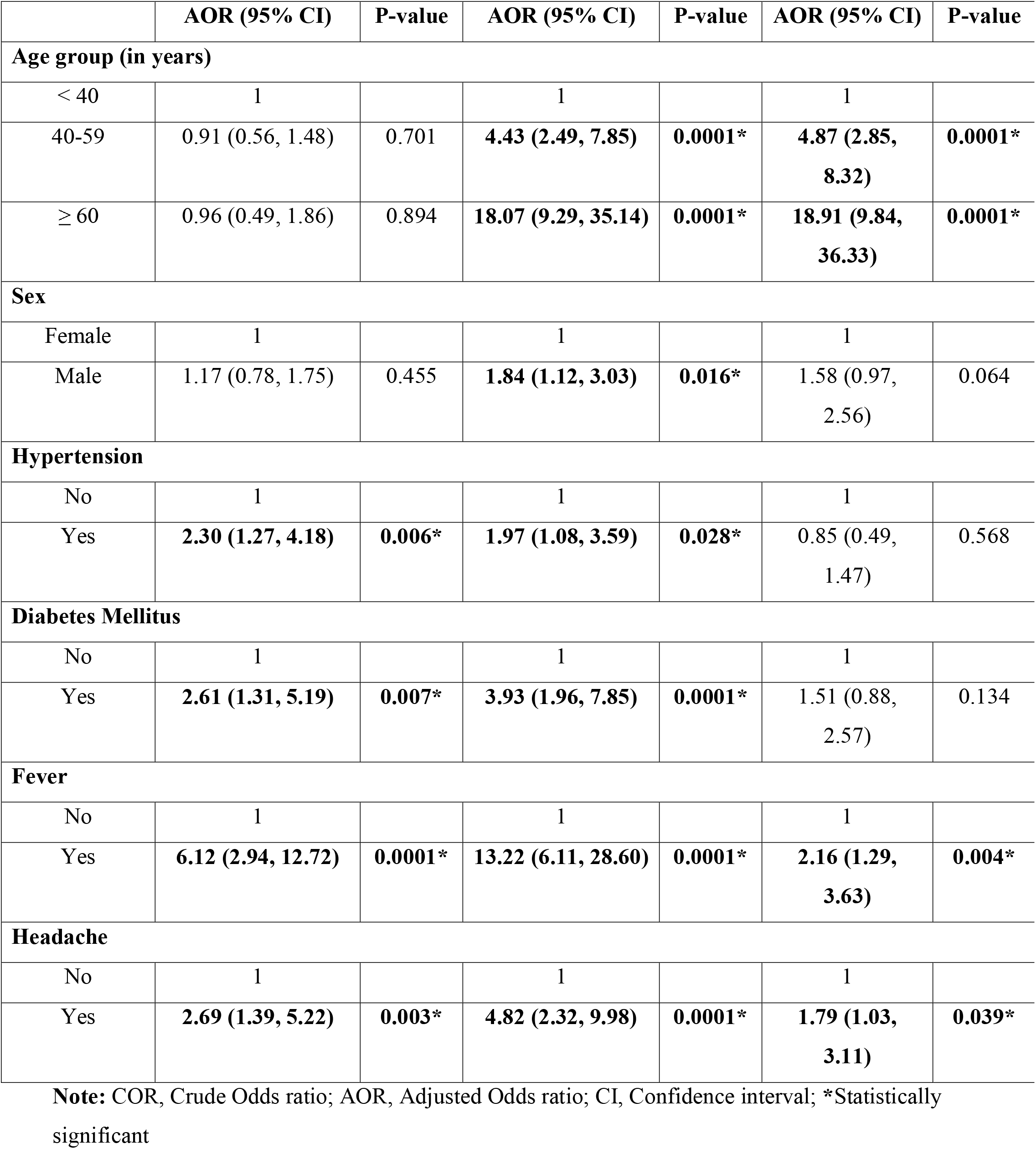
Results for the final multinomial logistic regression model among COVID-19 patients (n=686)

| Variable | Moderate (Vs Mild) | Severe (Vs Mild) | Severe (Vs Moderate) |
| --- | --- | --- | --- |

|  | AOR (95% CI) | P-value | AOR (95% CI) | P-value | AOR (95% CI) | P-value |
| --- | --- | --- | --- | --- | --- | --- |
| **Age group (in years)** |  |  |  |  |  |  |
| < 40 | 1 |  | 1 |  | 1 |  |
| 40-59 | 0.91 (0.56, 1.48) | 0.701 | **4.43 (2.49, 7.85)** | **0.0001\*** | **4.87 (2.85, 8.32)** | **0.0001\*** |
| ≥ 60 | 0.96 (0.49, 1.86) | 0.894 | **18.07 (9.29, 35.14)** | **0.0001\*** | **18.91 (9.84, 36.33)** | **0.0001\*** |
| **Sex** |  |  |  |  |  |  |
| Female | 1 |  | 1 |  | 1 |  |
| Male | 1.17 (0.78, 1.75) | 0.455 | **1.84 (1.12, 3.03)** | **0.016\*** | 1.58 (0.97, 2.56) | 0.064 |
| **Hypertension** |  |  |  |  |  |  |
| No | 1 |  | 1 |  | 1 |  |
| Yes | **2.30 (1.27, 4.18)** | **0.006\*** | **1.97 (1.08, 3.59)** | **0.028\*** | 0.85 (0.49, 1.47) | 0.568 |
| **Diabetes Mellitus** |  |  |  |  |  |  |
| No | 1 |  | 1 |  | 1 |  |
| Yes | **2.61 (1.31, 5.19)** | **0.007\*** | **3.93 (1.96, 7.85)** | **0.0001\*** | 1.51 (0.88, 2.57) | 0.134 |
| **Fever** |  |  |  |  |  |  |
| No | 1 |  | 1 |  | 1 |  |
| Yes | **6.12 (2.94, 12.72)** | **0.0001\*** | **13.22 (6.11, 28.60)** | **0.0001\*** | **2.16 (1.29, 3.63)** | **0.004\*** |
| **Headache** |  |  |  |  |  |  |
| No | 1 |  | 1 |  | 1 |  |
| Yes | **2.69 (1.39, 5.22)** | **0.003\*** | **4.82 (2.32, 9.98)** | **0.0001\*** | **1.79 (1.03, 3.11)** | **0.039\*** |

**Note:** COR, Crude Odds ratio; AOR, Adjusted Odds ratio; CI, Confidence interval; \*Statistically significant

## Discussion

Based on our findings from the 1000 bed makeshift hospital in Addis Ababa, risk factors associated with severe COVID disease on the multivariable multinomial logistic regression at 5% level of significance were age group, hypertension, diabetes mellitus, fever, and headache. Age group was one of the identified significant risk factors of disease severity. For patients in the age range of 40 to 59 years and 60 years and older, the odds of having severe disease as compared with mild and moderate disease was found to be higher compared with those patients younger than 40 years. But age group didn’t show a significant association with disease severity between moderate and mild cases. That means patients 40 years and above are at risk of developing more severe disease with the risk being much higher (18 fold) for those 60 years and above. This could be associated with the vulnerable nature of old age group due to the natural diminishing of the body’s defense mechanism and also the increased possibility of having concomitant comorbid illnesses that might not even be diagnosed, especially in the developing world with inadequate screening services, that further compromises the immune system. In another study conducted in our Center, older patients were found to be at risk of developing symptomatic infection compared to the younger group showing that older patients are susceptible to have a worse disease presentation and severity that could lead to a worse prognosis [16]. In addition, studies conducted in China also showed that aged people are more susceptible to development of severe disease and unfavorable outcome [9, 12, 17, 18].Being male was associated with a 1.842 odds of having severe disease as compared with mild disease than females. This significant difference in disease severity could be attributed to the identified difference in the disease biochemical activity between the two sexes showing that Angiotensin-converting enzyme 2, the receptor used by SARS-CoV-2, is found to be naturally abundant among males making it more convenient for high viral replication and development of symptomatic and severe disease compared to females [19-25].

For patients with hypertension and diabetes, the odds of having moderate and severe disease as compared with mild disease were higher compared to patients with no such illnesses. As explained above, since having one or more co-morbid illness results in a decreased immune defense mechanism of the body, it increases the patients’ probability of developing a disease from any infectious agent. This effect is accelerated if the comorbid illness/s is not well controlled. Furthermore, patients with comorbidity tend to be older, that in-turn adds to the existing decrease in immunity. This finding is also supported by other studies conducted in China, England and US [9, 12, 17, 18].

In addition, the other important factors that determine disease severity were fever and headache. The odds of having moderate and severe disease as compared with mild disease and also the odds of having severe disease as compared with moderate disease for patients with fever and headache were higher than patients with no such symptoms.. That means, having symptoms from COVID-19 infection are associated with developing a more severe disease as compared to the asymptomatic patients. Though these symptoms are not directly related to the disease severity classification in the study set up; like symptoms of cough, shortness of breath, and chest pain, they are found to be significant risk factors of disease severity. This implies that non-respiratory symptoms also might have a predictive value in disease categorization.

When interpreting these findings, it is important to consider both its strength and limitation. Its strength is that it included a robust sample size from the most representative care centre in the country. Its main limitation was that, based on prior reports, important variables that were potential risk factors for disease severity were not consistently available for all patients so could not be considered in the final model. This included Body Mass Index (BMI), as well as some laboratory and radiologic data.

## Conclusion

In Ethiopia, being 40 years and older (especially those 60 years and older), male, with a diagnosis of hypertension or diabetes, and having fever and headache were associated with severe COVID-19 disease. This suggests the initial cases in Ethiopia may have been milder because it affected younger people with fewer risk factors. We recommend a multicenter study that includes additional clinical, laboratory and radiologic data to further understand COVID-19 in Ethiopia.

## Data Availability

All relevant data are available upon reasonable request

## List of Abbreviations

CI: Confidence Interval
COVID-19: Coronavirus Disease 2019
FiO2: Fraction of Inspired Oxygen
LDH: Lactate Dehydrogenase
OR: Odds Ratio
PaO2: Partial Pressure of Oxygen
SARS-COV-2: Severe Acute Respiratory Syndrome Coronavirus 2
SpO2: Saturation of Oxygen
RT-PCR…: Real Time Polymerase Chain Reaction

## Authors Contribution

All authors contributed to the conception of the study. TWL designed the study, revised data extraction sheet, performed statistical analysis, and drafted the initial manuscript. All authors obtained patient data. All authors revised the manuscript and approved the final version.

## Acknowledgment

The authors would like to thank St. Paul’s Hospital Millennium Medical College for facilitating the research work.

## Availability of data and materials

All relevant data are within the manuscript and its Supporting Information files.

